# Leveraging Clinical, Functional, Molecular and Population Genetic Data Reveals Genotype Phenotype Association and Health Disparity in a Monogenic Disorder, CTX

**DOI:** 10.1101/2024.04.15.24305853

**Authors:** Jennifer Hanson, Penelope E. Bonnen

**Author notes:** Correspondence to: Dr. Penelope E Bonnen.

## Abstract

Cerebrotendinous Xanthomatosis (CTX) is a lipid storage disease caused by recessively inherited pathogenic variants in *CYP27A1* (OMIM 213700). The classic clinical presentation includes infantile-onset chronic diarrhea, juvenile-onset bilateral cataracts, with development of tendon xanthomas and progressive neurological dysfunction. These multisystem clinical features typically appear in different decades of life often confounding diagnosis of CTX. Further complicating diagnosis is the generally held belief that the clinical presentation of CTX varies highly between individuals and even within families. CTX is a treatable disorder and treatment is most effective when started in the first two decades of life, rendering a particular urgency to diagnosis.

In this study we bring a novel approach to detecting genotype phenotype associations in CTX. We conducted a systematic review of the literature to identify all functional analyses of pathogenic *CYP27A1* variants at the level of mRNA, protein and enzyme activity. We identified missense variants that result in complete loss of function (LOF) as well as missense variants that are have some partial function (hypomorphs). Next, we identified every CTX patient in the medical literature whose genotype and clinical phenotype were reported, and binned them according to functional genotype: LOF vs hypomorph. Analysis of these clinical, biochemical and molecular genetics data revealed a clear genotype phenotype association for CTX based on individuals who had two LOF variants vs two hypomorphs. The prevalence of each clinical feature was significantly higher in individuals with two LOF variants for every feature except tendon xanthoma and pyramidal signs. CTX had a detrimental effect on cognition for almost everyone with two LOF variants (96%), while tendon xanthomas were the most common feature in individuals with two hypomorphs (88%). We suspect this is due to ascertainment bias; individuals with a milder form of CTX may not get diagnosed with CTX unless they have this unusual hallmark of the disease. We studied the population genetics of the pathogenic *CYP27A1* alleles in gnomAD (N∼800,000). Estimated disease incidence based on carrier frequencies was consistent across the African/African American, Admixed American and European populations (1/308,000). However, no African/African American individuals have been reported in the medical literature as having CTX. Analyses of the pathogenic alleles in each population showed that the frequency of hypomorph pathogenic *CYP27A1* alleles was twice as high in African/African Americans (p=3.6E-4) vs Europeans (p=1.2E-4). Conversely, LOF alleles had a lower frequency in African/African Americans than in Europeans, p=6.1E-4 vs p=8.6E-4, respectively. By combining clinical, molecular, functional and populations genetics we uncovered a large health disparity in the diagnosis and treatment of CTX in African Americans and point to the milder clinical presentation of hypomorphs as an underlying component. The results of this study reveal specific opportunities for mitigating this disparity through recognition of the milder form of CTX as a clinical entity that is driven by hypomorph genetic alleles and broad adoption of biochemical testing that utilizes more sensitive biomarkers. Applying the framework and concepts leveraged in this study to the diagnosis of all monogenic disorders will likely result in improved diagnosis and health equity for the rare disease community.

**Key findings:**

- Joint analysis of clinical, functional, molecular, and population genetic data reveals health disparity in African Americans in a rare monogenic disorder, CTX.
- The gene that causes CTX, *CYP27A1*, harbors pathogenic missense variants that are loss of function and other pathogenic missense variants that are hypomorphs.
- Genotype phenotype analyses based on functional genotype - loss of function vs hypomorph - revealed a phenotype x functional genotype association for CTX.
- Individuals with loss of function genotype have a significantly more severe clinical presentation than those with a hypomorph genotype.
- Nearly all individuals with CTX who have a loss of function genotype have detrimental effects to their cognition (96%). The only exceptions to this received treatment with CDCA in the first decade of life.
- Population genetic analyses estimate that incidence of CTX is consistent across Blacks and Whites but systematic review of the medical literature returned no Black individuals having been reported to have CTX.
- Hypomorph pathogenic variants in *CYP27A1* occur more frequently in African/African Americans (p=3.6E-4) than Europeans (p=1.2E-4). The milder clinical presentation of the hypomorph genotype likely contributes to the under-diagnosis and misdiagnosis of African/African Americans with CTX.

## Introduction

Cerebrotendinous Xanthomatosis (CTX) is an inborn error of metabolism the hallmark features of which include chronic diarrhea, juvenile-onset cataracts, tendinous xanthomas and progressive neurological dysfunction (OMIM 213700). CTX is caused by recessive variants in *CYP27A1* which encodes for the enzyme sterol 27-hydroxylase which functions in bile acids synthesis. Bi-allelic pathogenic variants in this gene result in decreased cholic acid and chenodeoxycholic acid synthesis and consequently cause elevations of cholestanol, bile acid intermediates and bile alcohols.

Diagnosis can be made with clinical diagnostic testing of plasma cholestanol, bile acid intermediate and bile alcohol levels, urinary bile alcohol levels and/or genetic testing of *CYP27A1*. While some subjects experience symptoms from infancy, diagnosis is often not made until the third or fourth decade of life due to the multisystem clinical presentation and the emergence of features over decades (1, 2). Treatment with chenodeoxycholic acid (CDCA) is available that significantly slows progression of disease and when started early can prevent the major clinical problems associated with this disease, underscoring the importance of early diagnosis (1, 3).

Many reports assert that CTX clinical presentation is highly variable across individuals, even within families (2, 4–7). As clinical genetic testing has surged in recent years, individuals with CTX are being identified at younger ages; understanding the natural history of CTX and any potential genotype phenotype associations could potentially benefit patients and families. However, prior studies report there is no genotype phenotype association in CTX.

In this study we brought a novel approach to detecting genotype phenotype associations in CTX and we leveraged those findings alongside population genetic analyses to identify health disparities. We conducted a systematic review of the literature to identify every CTX patient in the literature who had a genotype and clinical phenotype described. We conducted a systematic review of the literature to identify all instances of functional analyses of pathogenic CTX variants including mRNA, protein and enzyme activity. Through this effort we identified *CYP27A1* missense variants that result in loss of function (LOF) as well as missense variants that are hypomorphs (have some partial function). Every CTX patient was grouped according to functional genotype: LOF vs hypomorph and tested for genotype phenotype association. We also studied the population genetics of *CYP27A1* variants with attention to their functional consequence. By leveraging this multi-modal dataset including clinical, functional, molecular genetic and population genetic data we discovered a genotype phenotype association for CTX in LOF vs hypomorph variants and show that the milder clinical phenotype of individuals with CTX who have hypomorphs may underlie a large health disparity in the diagnosis and treatment of African Americans with CTX.

## Results

### Genotype Phenotype Correlation for CYP27A1 LOF Genotype

Clinical diagnostic criteria refer to cardinal features of CTX: cataracts, tendon xanthomas, cerebellar signs, pyramidal signs, peripheral neuropathy, chronic diarrhea (1, 2). However, many studies report that CTX clinical presentation is highly variable across individuals, even within families (2, 4–7). No genotype-phenotype studies have been conducted that reported a robust sample size or statistical analysis. There does not appear to be a sufficient number of samples with the same genotype for powerful statistical analysis. For example, Verrips, *et al.* 2000 conducted a comparison across individuals who were homozygous for the same *CYP27A1* pathogenic variant; in 79 individuals who had a homozygous genotype there were 23 different variants. (4).

In light of this, we utilized a function-based paradigm to query if there was a genotype phenotype association for CTX: a study that compares the clinical presentation of individuals who possess two LOF *CYP27A1* variants versus individuals who possess two hypomorph missense variants. This function-based paradigm requires that LOF variants do not possess any enzyme activity; likewise, hypomorph variants must confer partial enzyme activity. We conducted a review of the literature to find studies that reported the resulting effect of pathogenic *CYP27A1* variants on mRNA, protein or enzyme activity and identified *CYP27A1* variants that cause CTX that show partial enzyme activity and those that result in complete loss of enzyme activity.

There are reports of nonsense, splice or frameshift *CYP27A1* variants that result in no enzyme activity, stable mRNA or protein (8–16). These are LOF alleles. Interestingly, there are several missense variants that have been demonstrated to be LOF variants. These missense variants have been functionally vetted in patient fibroblasts or through *in vitro* expression assays and been shown to result in no stable protein and/or no enzyme activity (12, 13, 17–28). These can be described as falling into three categories: missense variants that are physically present in the last amino acid of an exon and have been demonstrated to affect splicing and result in no enzyme activity; missense variants that are physically present in the middle of an exon and have been demonstrated to affect splicing and result in no enzyme activity; and missense variants that do not affect spicing but have been demonstrated to result in no enzyme activity.

Missense variants that affect the last amino acid in an exon and result in aberrant splicing are: c.1183C>T;p.(Arg395Cys) (26, 28), c.1184G>A;p.(Arg395His) (21), c.1183C>A;p.(Arg395Ser) (19), c.646G>C;p.(Ala216Pro) (12, 13, 25). An additional variant at the same amino acid that does not have published functional data but is bioinformatically predicted to change splicing is c.646G>A;p.(Ala216Thr) (SpliceAI = .5 donor loss, Pangolin =.99 splice loss). In addition, pathogenic variant, c.1016C>T;p.(Thr339Met), affects the last amino acid of exon 5 and was demonstrated to result in loss of enzyme activity even though it is not bioinformatically predicted to affect splicing (24). Missense variants that affect splicing but are not at the boundaries of an exon include c.435G>T;p.(Gly145Gly) (18) and c.434G>C;p.(Gly145Ala) (24). There are pathogenic missense variants that are internal to an exon for which we did not find any functional studies, but bioinformatics analyses predicted splice gain: c.667G>C;p.(Glu223Gln) (SpliceAI = .87, Pangolin = .80); c.776A>G;p.(Lys259Arg) (SpliceAI =

.98, Pangolin = .72); c.804G>T;p.(Trp268Cys) (SpliceAI = .84, Pangolin = .34); c.1298G>A; p.(Arg433Gln) (SpliceAI .25, Pangolin = 0.03). Missense variants that resulted in undetectable levels of protein and enzyme activity in patient fibroblasts but do not appear to affect splicing include: c.1421G>A;p.(Arg474Gln) (20, 22, 23), c.1214G>A;p.(Arg405Gln) (20, 23), c.1213C>G;p.(Arg405Gly) (25). The missense variant affecting the same amino acid c.1213C>T;p.(Arg405Trp), does not have published functional data but was excluded from our analysis of patients with missense variants.

Pathogenic hypomorphs have also been reported for *CYP27A1*. Missense variants in *CYP27A1* that cause CTX have been demonstrated to have full or partial protein or mRNA levels alongside partial enzyme activity in patient fibroblasts or *in vitro* expressions systems (22–26, 29, 30). These missense variants are therefore be classified as hypomorphs.

For genotype phenotype analyses individuals with CTX were binned into two groups. Those who had two LOF variants, defined as nonsense or frameshift variants, were grouped together and termed LOF. Those individuals who had two missense variants that were functionally demonstrated to have partial enzyme function were grouped together and referred to as hypomorphs. Individuals who had missense variants that were demonstrated to result in LOF were excluded from this analysis. Additionally, individuals who had splice variants were excluded from this analysis.

### Individuals with a LOF genotype have a more severe CTX phenotype

Individuals with CTX who had two LOF variants were identified and their clinical presentation was assessed (N=67). The frequency of clinical features in the LOF group was: cataracts 90%, tendon xanthomas 64%, cognition deficits 96%, cerebellar signs 81%, pyramidal signs 72%, peripheral neuropathy 36%, parkinsonism 20%, diarrhea 28% (Figure 3). By comparison, individuals with two hypomorph missense variants (N=43) showed clinical features in different frequencies: cataracts 42%, tendon xanthomas 88%, cognition deficits 60%, cerebellar signs 58%, pyramidal signs 74%, peripheral neuropathy 14%, parkinsonism 2%, diarrhea 14% (Figure 3). The distribution of age at last report was significantly different between these two groups with more LOF individuals at younger ages and more hypomorphs at the other end of the age distribution (Figure 2). The prevalence of each clinical feature was significantly higher in individuals with two LOF variants for every feature except tendon xanthoma and pyramidal signs. This is noteworthy in light of the fact that the hypomorph cohort has older individuals in it giving them more time to manifest clinical features of CTX. Tendon xanthomas were the most common feature in individuals with two hypomorph variants. We suspect this is due to ascertainment bias; individuals with a milder form of CTX may not get diagnosed with CTX unless they have this unusual hallmark of the disease. The fact that pyramidal signs were found in ∼73% of CTX patients regardless of functional genotype renders this an important diagnostic sign of this disease.

**Figure 1.**
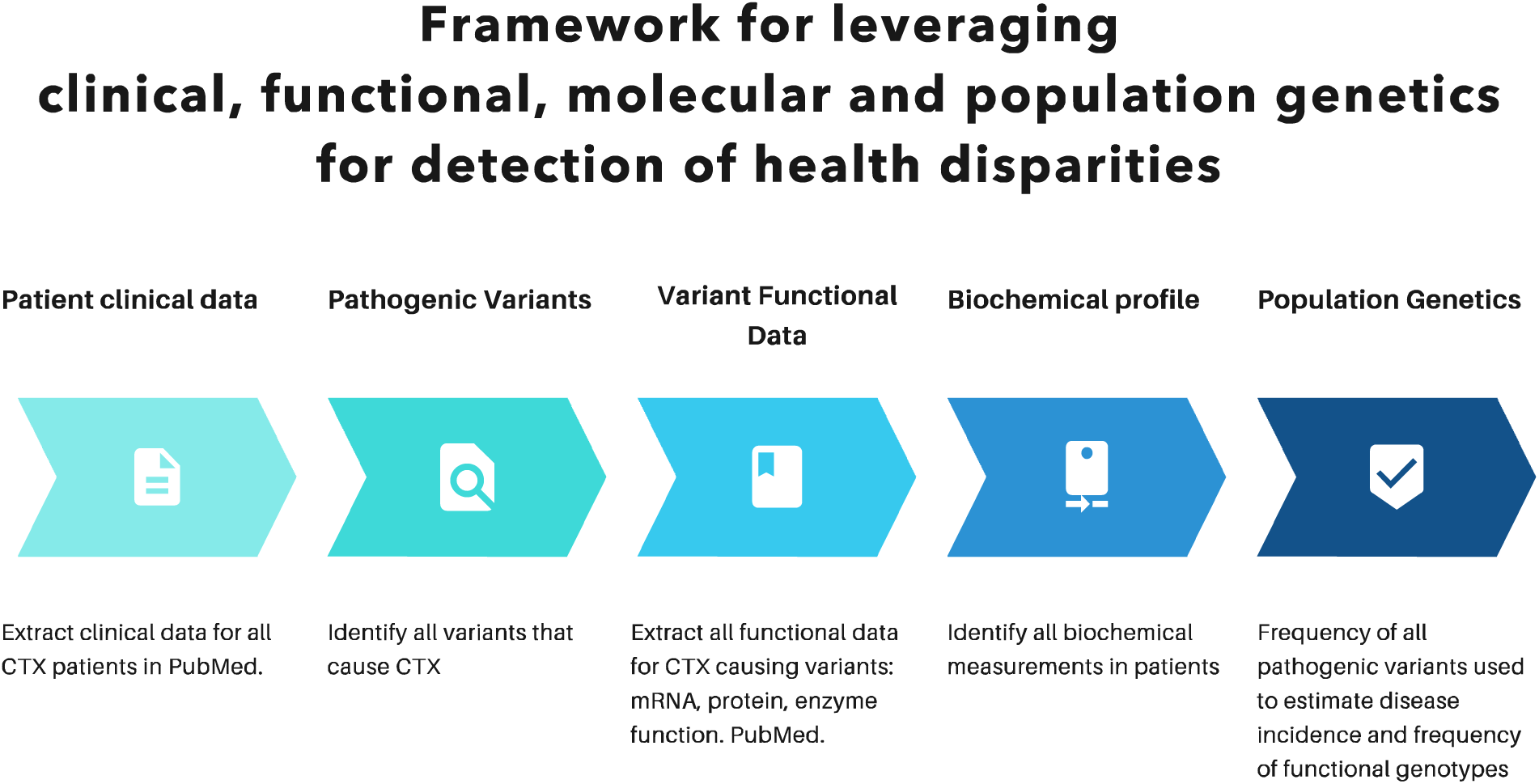
Analytical framework for leveraging multi-modal data types to identify genotype phenotype associations and health disparities.

**Figure 2.**
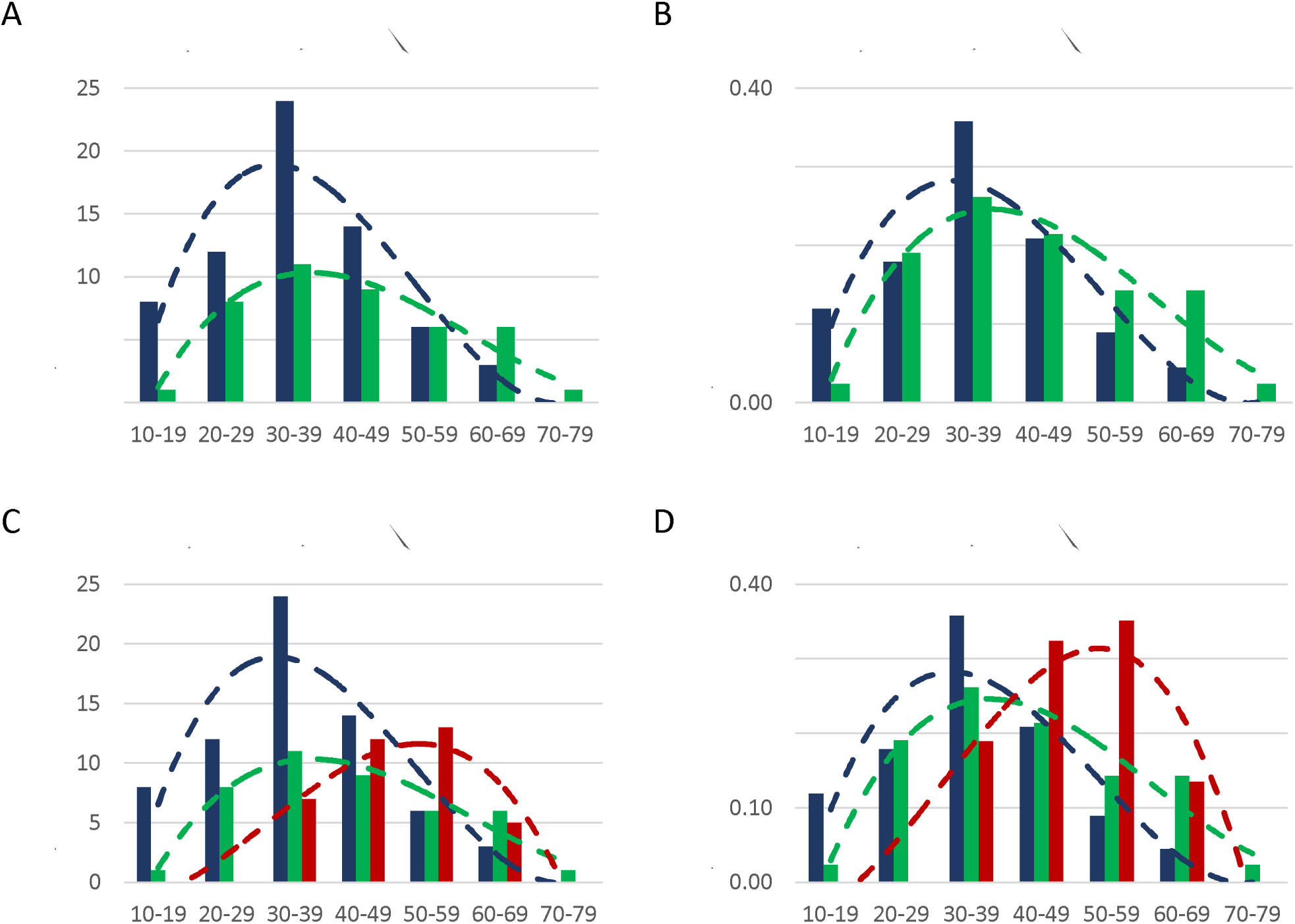
Age at Last Report for Individuals with CTX included in this study. Individuals with CTX who have two LOF *CYP27A1* variants are shown in blue. Individuals with CTX who have two hypomorph *CYP27A1* variants are shown in green. Individuals with CTX who have parkinsonism, regardless of genotype, are shown in red.

**Figure 3.**
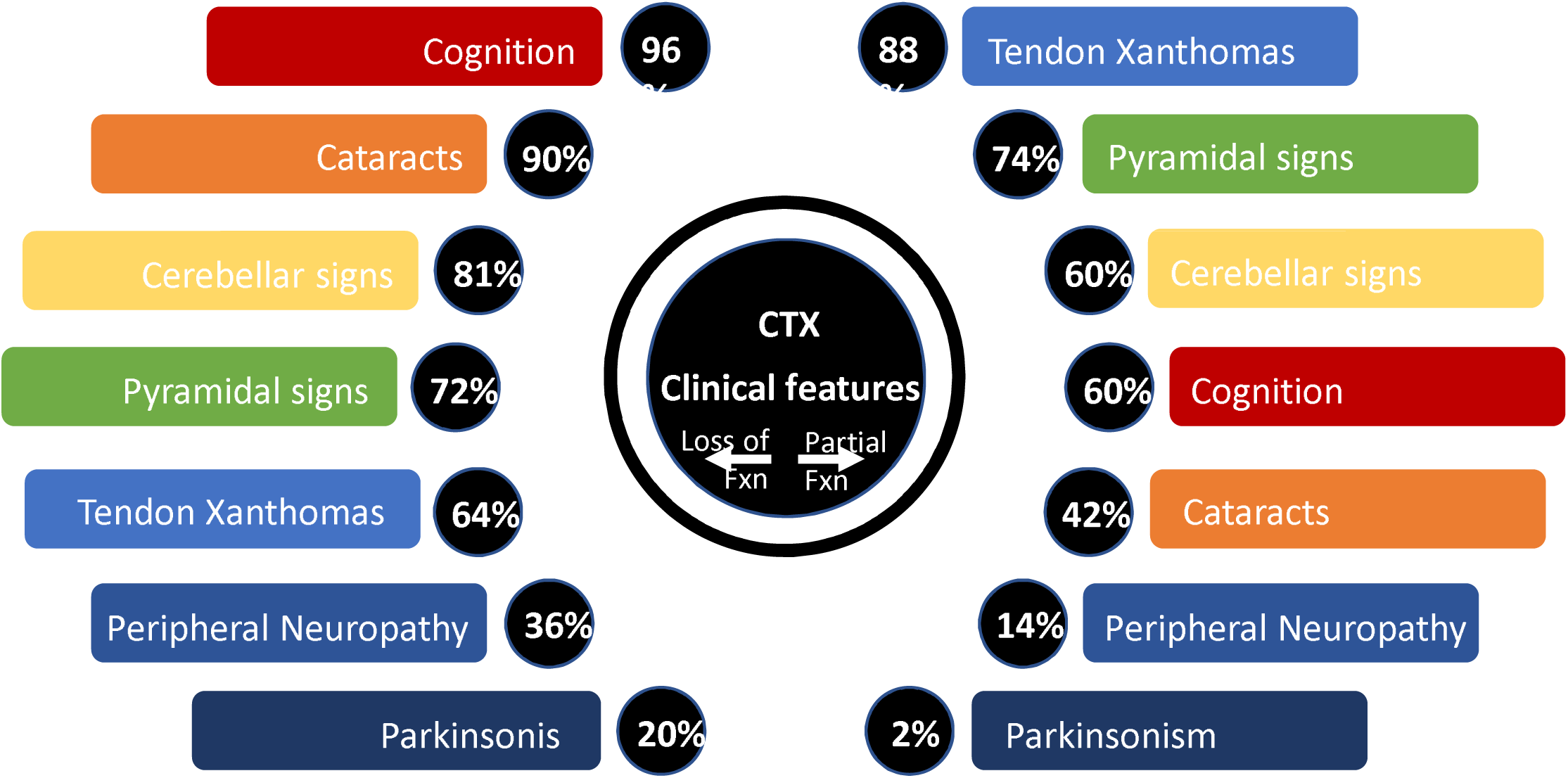
Clinical Features of Individuals with CTX who have LOF vs hypomorph genotypes. Individuals who have two LOF *CYP27A1* variants were grouped together and the frequency of each clinical feature of CTX in that group is shown here in the left side of the figure. Individuals who have two hypomorph *CYP27A1* variants were grouped together and the frequency of each clinical feature of CTX in that group is shown in the right side of the figure.

### CTX affects cognition in nearly all subjects with two LOF variants

In the context of CTX, cognitive deficits and cognitive decline may occur in childhood or adulthood, and depending on age of onset and severity will be described differently. The term intellectual disability (ID) is defined in the DSM-5 as significant cognitive deficits resulting in lowered ability in intellectual functioning and adaptive behaviors with an age of onset beginning in childhood. It can be diagnosed through standardized IQ testing resulting in less than 70 appearing before the age of 22. In contrast, dementia, which was named major neurocognitive disorder (NCD) in the DSM-5, is characterized by acquired cognitive deficits in adulthood, which represent a significant decline from prior level of functioning, rather than cognitive deficits present from childhood. In this study “cognition” was a composite of intellectual disability, learning difficulty, dementia or cognitive decline; if a person was reported to have any one of these terms we noted them as having effects on “cognition”.

CTX had an effect on cognition for almost everyone with two LOF variants (96%), while the 60% of the missense group had effects on cognition. The LOF group had intellectual disability 43%, learning difficulty 22%, dementia 36% or cognitive decline 66%; while the hypomorph group had intellectual disability 14%, learning difficulty 33%, dementia 16% or cognitive decline 30%. It’s striking that the incidence of ID is 3x higher in the LOF group than the hypomorph group. This illustrates the severity of the LOF phenotype and underscores the importance of early treatment for CTX.

### Parkinsonism is more frequent in CTX LOF patients

Parkinsonism was present in 20% (14/67) of LOF patients and 2% (1/43) of hypomorph patients. Parkinsonism is one of the less frequent manifestations of CTX and as a result the numbers were not large (LOF parkinsonism N=14, hypomorph parkinsonism N=1).

As an alternate means of testing if parkinsonism truly varied by functional genotype, we identified every CTX patient reported with parkinsonism. We defined parkinsonism as having two or more of the following: resting tremor, postural tremor, bradykinesia, rigidity, hypomimia and dystonia. We found 37 individuals in the literature reported as having CTX and parkinsonism (1, 2, 4, 16, 27, 31–65). The age distribution of this group of patients was older than the LOF or hypomorph patients: most individuals with parkinsonism were 50-59 years at last report (Figure 2). Of these 37 subjects with parkinsonism the genotype of 35 could be unambiguously determined to have LOF variants or not; two individuals were compound heterozygous for a splice variant and p.(Thr422Ala), a missense variant for which no functional data has been published. 91% (32/35) of subjects with CTX and parkinsonism had at least one LOF variant; 83% (29/35) had two LOF variants (frameshift, nonsense, canonical splice site variants or missense variants that have been functionally demonstrated to be LOF). Three individuals were compound heterozygous for a LOF variant and a missense partial function variant. Three subjects were homozygous for a missense hypomorph; one was homozygous for p.(Arg127Gln) and two were homozygous for p.(Arg474Trp).

### CYP27A1 Functional Genotype and Biochemical Phenotype

As an inborn error of metabolism, the gold standard for diagnosis of CTX is biochemical testing. 5α-cholestanol has historically been the most commonly used biochemical marker. No genotype phenotype association was reported between the genotype of a person with CTX and the cholestanol level of a person with CTX, and cholestanol is not utilized as a quantitative marker for the clinical severity of disease. In recent years clinical genetic testing has surged and new biochemical markers have been developed for testing of CTX including 5β-cholestane- 3α,7α,12α,25-tetrol-3-O-β-d-glucuronide (25-tetrol glucuronide), 5β-cholestane- 3α,7α,12α,23S,25-pentol (23S-pentol), 7α–hydroxy-4-cholesten-3-one (aka 7αC4 or 7aHCO) and 7α,12α-dihydroxy-4-cholesten-3-one (aka 7α12αC4 or 7α12α-diHCO) (30, 66–68). These markers appear to be more sensitive than cholestanol and their use has led to reports of individuals with CTX who have elevations in these markers while having normal or slightly above normal cholestanol (30, 69–76). These individuals were noted to have a milder clinical course of CTX.

The genotypes of all individuals in the literature reported to have a genetically confirmed diagnosis of CTX and normal or high normal cholestanol levels when not taking medication (CDCA/Cholic acid) (N=10) were examined (30, 69–76). The range in age at last report for this group of individuals was 29-86 years. All ten individuals were reported to have tendon xanthomas, without which they most likely would not have been diagnosed with CTX. The second most common features were dementia and pyramidal signs, with 30% (3/10) displaying these features. Two subjects had gait abnormality or ataxia. The genotypes of these ten individuals showed that no one had a LOF genotype. Three individuals were homozygous for two hypomorphs. One subject was compound heterozygous for a LOF variant and a hypomorph variant. Three more subjects were compound heterozygous for a LOF variant and a missense variant for which we have no functional data. Three additional individuals had missense variants that were not canonical LOF variants but there was no functional data for those variants.

### Population Genetics x Functional Genotypes Points to Under-diagnosis of CTX in African Americans

Given the results of this study showing that LOF versus hypomorph variants result in more severe CTX clinical presentation we asked what *CYP27A1* variants were most frequent in global populations. We queried the gnomAD population database of ∼800,000 individuals and asked the frequency of known pathogenic *CYP27A1* variants in the African/African American, Admixed American, Asian and European populations. We summed the frequency of all pathogenic variants in a population and used that frequency to calculate incidence according to Hardy Weinberg principles. Using the carrier allele frequency of pathogenic variant in a population resulted in an incidence of CTX 1/300,000 in African, Admixed American and European populations. Asians had a higher incidence of CTX, 1/80,000 (Figure 4). Despite this seemingly consistent incidence of CTX across African/African American, Admixed American and European populations, search of the medical literature did not return a single person with CTX being reported as being Black, African American, African or being from any African-derived population.

**Figure 4.**
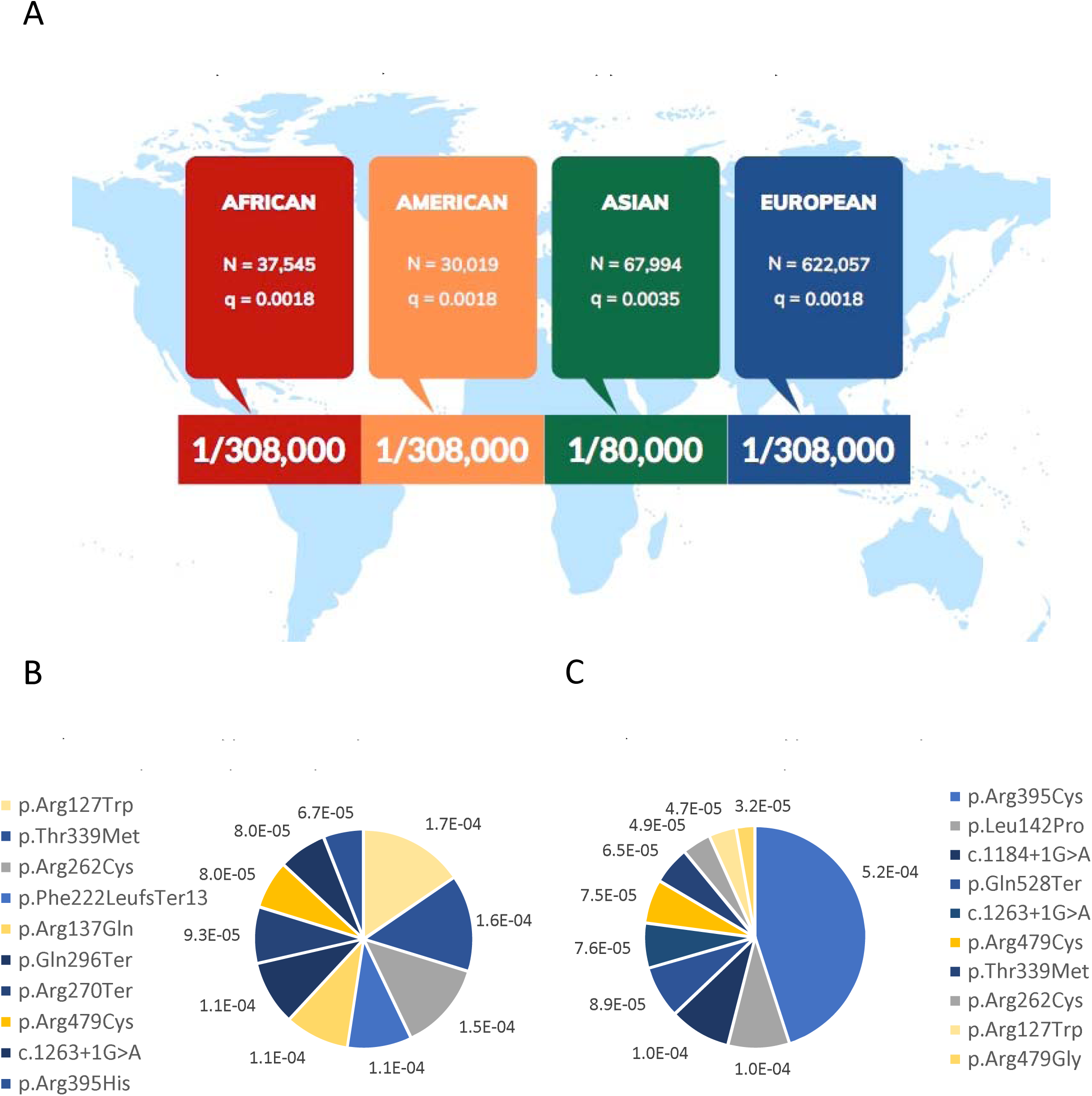
Population genetics approach to identifying health disparities. A. The incidence of CTX was estimated in four global populations by applying Hardy Weinberg principle to carrier allele frequencies for pathogenic CTX variants in *CYP27A1*. N=the number of individuals in each population who was included in the allele frequency calculation. q is the sum of the allele frequencies of all pathogenic variants in a population. Incidence estimates are shown as 1/X number of individuals. B. The most common CTX pathogenic alleles in the African/African American population in gnomAD is shown with hypomorphs colored yellow and LOF shaded blue. Variants for which no functional information was available are shaded grey. C. The most common CTX pathogenic alleles in the European population in gnomAD is shown with hypomorphs colored yellow and LOF shaded blue. Variants for which no functional information was available are shaded grey.

We queried top ten most frequent *CYP27A1* pathogenic variants in gnomAD in the African/African American population. The most frequent variant in the African/African American population is c.379C>T;p.Arg127Trp with a frequency of 1.7E-4. This variant has been demonstrated to confer partial enzyme activity (22, 23, 29). This variant is found in other populations: Admixed Americans 7E-5, Europeans 5E-5, and Asians 3.7E-5. A number of patients have been published with this variant, most of them appear to have a milder clinical presentation, including two who had normal cholestanol (4, 30, 38, 56, 69, 74, 75, 77–83).

The second most frequent pathogenic variant in *CYP27A1* in the African population is c.1016C>T;p.Thr339Met, 1.6E-4. This missense variant was reported to display no enzyme activity *in vitro* with enzyme activity measured as D3 25-hydroxylase activity (24). This variant was first reported in Jewish CTX patients and is found in gnomAD all over the globe: 1.5E-4 in Admixed Americans, 3.2E-5 in Asians, and 6.5E-5 in Europeans (9). Many CTX patients with this variant have been reported in the literature, however, none of them were noted to be Black.

The third highest variant in Africans/African Americans is c.784C>T;p.Arg262Cys, 1.5E-4. There is no functional data to indicate what enzyme activity level may be conferred by this variant. Bioinformatically, it is predicted to be pathogenic (Damaging by Sift and PolyPhen, CADD v1.7=27), but it is not predicted to result in LOF. There is a paucity of CTX patients reported to have this variant. It was reported in a Japanese patient who had no signs of disease until age 57 and was diagnosed at 61 with elevated cholestanol (84). The man presented with a rapidly progressing spasticity and sensory disturbance in the lower limbs. Spinal MRI showed long hyperintense lesions. No other signs of CTX were present. Individuals with this clinical presentation may not always receive a diagnosis of CTX.

All of the top ten most frequent *CYP27A1* pathogenic variants in gnomAD in the African/African American population were plotted by frequency and color coded for functional status (Figure 4). If a variant has been demonstrated to be complete loss of function it was colored blue and if a variant has been demonstrated to retain some enzyme activity it was colored in yellow. If no functional data was available for a variant it was colored in gray. The same was done for the top ten most frequent pathogenic variants in Europeans (Figure 4). In African/African Americans these ten variants combined have a frequency of 1.2E-3 and in Europeans the most frequency *CYP27A1* variants have a combined frequency of 1.1E-3. This comparison shows that the African/African American population has a larger proportion of partial function alleles than the European population. The combined frequency of partial functioning pathogenic *CYP27A1* alleles in African/African Americans is 3.6E-4 vs 1.2E-4 in Europe. Conversely, LOF alleles have a lower frequency in African/African Americans than in Europeans, 6.1E-4 vs 8.6E-4, respectively.

The most frequent *CYP27A1* pathogenic variant in Europeans is c.1183C>T;p.(Arg395Cys), p=5E-4. This was documented to result in no enzyme activity *in vitro* (85). It occurs in the last amino acid of exon 6 and is bioinformatically predicted to affect splicing. An alternate amino acid substitution at Arg395, c.1183C>A;p.(Arg395Ser), was demonstrated *in vitro* to affect splicing and result in no enzyme activity *in vitro* (19). Another substitution at Arg395, c.1184G>A;p.(Arg395His), is also bioinformatically predicted to affect splicing and was shown in patient fibroblasts to result in no enzyme activity (21).

Pathogenic variant, p.(Arg395Cys), is not only the most frequent pathogenic variant in Europe it is the most common *CYP27A1* pathogenic variant globally, according to gnomAD v4.0. The second most frequent variant globally is also loss of function: canonical splice variant c.1184+1G>A was demonstrated to affect splicing of *CYP27A1* in patient fibroblasts (13). Despite being the second most common pathogenic globally, it does not appear in African/African Americans in gnomAD v4.0.

In this study we report a genotype-phenotype association where individuals with two LOF variants have a more severe clinical presentation of CTX than those with two partial functioning pathogenic variants of *CYP27A1*. Based on this, and the fact that the African/African American population has a higher frequency of pathogenic partial functioning *CYP27A1* variants, we hypothesize that CTX may be more likely to exhibit a milder clinical presentation in this population and lead to under-diagnosis and misdiagnosis. These findings point to specific opportunities for identifying African American CTX patients by acknowledging that CTX as a clinical entity includes individuals with a milder phenotype driven by hypomorph alleles and the need for more sensitive biochemical testing.

## Discussion

The finding of a genotype phenotype association for CTX is a paradigm change. Several studies have reported no genotype phenotype association for this disease including often stating that clinical presentation even varies within families. However, these studies lacked sufficient sample size for a strict genotype-phenotype association study and often did not report statistical analyses. In the context of rare disease, it is difficult to achieve large enough numbers of individuals with the same genotype to have sufficient statistical power. We took a different approach for CTX and asked if individuals with two LOF variants had a different phenotype than individuals with two variants that have partial enzyme activity (hypomorphs). This binning of subjects enabled us to increase sample size, and therefore power, and is based on a biological foundation. This is the first time that the body of literature that reported functional testing of *CYP27A1* variants in patient fibroblasts and *in vitro* was taken in to account when considering genotype phenotype associations for CTX. A particularly salient and important aspect of the genetics of CTX is that there are several *CYP27A1* missense variants that have been demonstrated to be LOF variants and some have partial enzyme function. This key factor, that some missense variants have residual enzyme activity and others do not, enabled us to discern that individuals with CTX who have two variants that confer partial enzyme activity have a milder clinical presentation and are more likely to have normal cholestanol levels.

Simply categorizing CTX patients into two groups: those individuals with two LOF variants and those individuals with two hypomorph variants revealed a clear distinction in CTX clinical presentation. Those with two LOF variants (N=67) had a more severe clinical course: cataracts 90%, tendon xanthomas 64%, cognition deficits 96%, cerebellar signs 81%, pyramidal signs 72%, peripheral neuropathy 36%, parkinsonism 20%, diarrhea 28%. By comparison, individuals with two hypomorph missense variants (N=43) had a milder phenotype and showed clinical features in different frequencies: cataracts 42%, tendon xanthomas 88%, cognition deficits 60%, cerebellar signs 58%, pyramidal signs 74%, peripheral neuropathy 14%, parkinsonism 2%, diarrhea 14%. The age at last report was older in the hypomorph cohort giving them more time to manifest clinical features of CTX and yet they showed fewer signs of disease. It stands to reason that a patient with residual enzyme activity would experience a milder phenotype. This has been documented for other inborn errors of metabolism including Gaucher disease, phenylketonuria and Pompe disease.

CTX had an effect on cognition for almost everyone with two LOF variants (96%). This group of subjects experienced intellectual disability (43%), learning difficulty (22%), dementia (36%) and/or cognitive decline (66%). These data are extracted from the literature and while we applied a systematic use of terms to every subject, we did not clinically examine every subject and there are almost certainly some aspects that are not fully ascertained. For example, IQ testing was not conducted on every person reported in the literature as having CTX; but in this study a subject was categorized as having ID only if IQ testing was performed before the age of 22 and showed an IQ of less than 70. Similarly, an MMSE was not conducted on every subject. In the absence of consistent testing on every subject, ascertainment is likely incomplete. And yet, even in the face of incomplete ascertainment it is remarkable to find that nearly every subject with two LOF variants suffered some effects on cognition. In fact, two of the three individuals in this study who had two LOF variants but did not display some effect on cognition received early treatment with CDCA: one was started on CDCA at age 2 and the other at age 7. The CDCA likely prevented the disease from affecting their cognition and underscores the benefits and importance of treatment for CTX. It also tells us that in the absence of treatment, all individuals with two LOF variants in *CYP27A1* would have cognitive deficits.

Consideration of the effects of CTX on cognition in milder cases is also important. A large proportion of the group of individuals with two hypomorphs (60%) had effects on cognition. This group experienced a much lower rate of intellectual disability (14%) than in the LOF group, and yet a higher proportion of the hypomorph group was noted to have 33% learning difficulty. It is possible that having CTX could result in a lower IQ than a person would have had otherwise. For example, an average person has an IQ of 100; if an average person has a milder case of CTX (hypomorphs) their IQ may lower to 90 whereas if they had a severe case of CTX (LOF) their IQ may lower to 65. The person with CTX and an IQ of 90 would not be diagnosed with ID and it may not be appreciated that CTX had any effect on their cognition; but that does not mean that CTX did not affect their cognition. This underscores the importance of early treatment for CTX for all cases.

Parkinsonism is one of the less common features of CTX and we showed through two different analyses that it happens significantly more often in individuals who have two LOF variants. In the first analysis in this study we observed that parkinsonism is more frequent in CTX LOF patients 20% (14/67) than in individuals with CTX who have two hypomorphs 2% (1/43). Next, we identified every person in PubMed reported to have CTX and parkinsonism (N=37). 91% of subjects with CTX and parkinsonism had at least one LOF variant; most of these had two LOF variants (frameshift, nonsense, canonical splice site variants or missense variants that have been functionally demonstrated to be LOF). Three subjects were homozygous for a missense hypomorph: one was homozygous for p.(Arg127Gln) and two were homozygous for p.(Arg474Trp). These three individuals were molecularly diagnosed before the year 2000. All three were reported as being homozygous in the absence of consanguinity. It is possible that these subjects have a second variant that was undetected. However, it is also possible that being homozygous for the same hypomorph variant may confer a more severe phenotype than being compound heterozygous for two different hypomorph variants. For example, CYP27A1 Arg474 forms a hydrogen bond with the proprionate group of the heme D-ring and mutation of this Arginine results in altered heme binding (86). Enzyme activity was measured in the fibroblasts of a person with CTX who was homozygous for this variant and it showed 1.4% activity compared to controls (22). Fibroblasts from the father of this patient, who was heterozygous for this variant and did not have CTX, showed CYP27A1 enzyme activity of 10% of controls (22). One possible explanation is that being homozygous for this variant impeded heme binding more than if a person was heterozygous for this variant and another CTX causing variant that affected a different function of the protein.

Interestingly, cholestanol has been demonstrated to induce the fragmentation, aggregation and spreading of α-synuclein in the brain resulting in the degeneration of the nigrostriatal dopaminergic pathway and parkinson motor symptoms (65). 5α-Cholestanol was measured in post-mortem brain of an individual with CTX and showed that cholestanol was present in multiple regions of the CTX brain including the substantia nigra (30). The genotype of this person was not reported but they were described as having tendon xanthoma, cataracts, ataxia, and below normal intelligence (87). This subject died when they were 26 years old and they were not noted to have exhibited parkinsonism and yet their brain showed cholestanol in the substantia nigra (87).

The clinical course for individuals with two *CYP27A1* hypomorphs appears to be milder, and further study is warranted to delineate the clinical and biochemical profile of this group of CTX patients. Individuals with two hypomorphs have been reported to have lower cholestanol and higher serum 25-hydroxyvitamin D3 (25(OH)D3) concentrations (23, 30, 73). Zhang *et al* reported serum 25-hydroxyvitamin D3 (25(OH)D3) concentrations in individuals who had CTX and compared the 25(OH)D3 values between patients with LOF variants vs patients with missense mutations. CTX patients with splicing, nonsense or frameshift variants (N=6) had lower 25(OH)D3 levels than those with two missense variants (N=3) (Mean + SEM: 5.9 + 1.4 ng/ml vs. 15.7 ± 5.4 ng/ml; P=0.04) (23). This study grouped patients who had two missense variants separately from individuals who had LOF variants; these particular missense variants happened to be hypomorphs. Other studies have reported individuals who had CTX and had plasma cholestanol levels in the normal to high normal range. Several of these subjects had hypomorph alleles, however, not all of the variants in these individuals have been tested to elucidate their functional effects. Systematic functional analyses of all pathogenic *CYP27A1* missense variants is warranted. The sample size of these studies is small but their findings support the notion that subjects with two hypomorphs may have a milder biochemical and clinical phenotype. Further study into the biochemical phenotype of individuals with hypomorphs may be helpful for improved diagnosis of CTX.

Tendon xanthomas were the most common feature in individuals with two hypomorphs. Moreover, all ten individuals reported with normal serum cholestanol while not on treatment were reported to have tendon xanthomas, which was likely the only reason they were diagnosed with CTX; for half of these subjects, tendon xanthoma was the only clinical feature reported. Just three diseases manifest xanthomas: CTX, Sitosterolemia, and Homozygous Familial Hypercholesterolemia. It is possible that there is a form of CTX that presents with most of the time with tendon xanthoma and few if any other features. However, we suspect that ascertainment bias has played a role in the diagnosis of individuals with milder forms of CTX as mild cases may not get diagnosed with CTX unless they have this unusual hallmark of the disease. We hypothesize that there are also individuals with hypomorphs who have mild cases of CTX that do not include tendon xanthoma and go undiagnosed for CTX. Given the pervasive effects on cognition that we see in this study, dementia clinics may be an important place to look for these patients. Pyramidal signs were the only clinical feature that appeared in the same frequency in LOF and hypomorphs so this may be an important hallmark to help diagnose patients with CTX who do not have xanthomas. In addition, CTX is an IEM and as such an effective strategy for identifying patients is biochemical testing using biomarkers that are more sensitive than cholestanol (30, 69, 88).

Given the difference in clinical presentation for LOF vs hypomorph CTX we studied the population genetics of *CYP27A1* pathogenic variants in global populations with attention to the functional status of these variants. When CTX incidence is calculated using the carrier frequency for all *CYP27A1* pathogenic variants within a population consistent incidence is observed across African/African Americans, Admixed Americans and Europeans, 1/308,000. However, no African/African American individuals with CTX were found in the medical literature. This disparity was investigated by considering the functional status of pathogenic alleles in each population. Interestingly, the combined frequency of hypomorph pathogenic *CYP27A1* alleles in African/African Americans is 3.6E-4 vs 1.2E-4 in Europe. Conversely, LOF alleles have a lower frequency in African/African Americans than in Europeans, 6.1E-4 vs 8.6E-4, respectively. This population genetic analysis of pathogenic *CYP27A1* variants points to under-diagnosis of CTX in African Americans, possibly as a result of having a higher frequency of hypomorphs. This apparent disparity in diagnosis and consequentially treatment of CTX can be overcome with universal screening for this treatable disease. Better delineation of the clinical and biochemical phenotype of individuals with CTX who have hypomorphs alongside the broad adoption of biochemical testing with more sensitive biomarkers will assist in improved diagnosis and mitigating this disparity.

## Methods

### Literature Search

Searches of medical literature were conducted to identify individuals with confirmed *CYP27A1* genotype and clinical details. We systematically reviewed the literature for all cases of CTX published up to December 31, 2023, written in English and contained in the National Institute of Health PubMed database. The search was conducted using the terms “cerebrotendinous xanthomatosis” and “CYP27A1”. We reviewed all papers returned by this search; all 557 published manuscripts returned by this search were studied for genetic, clinical and functional data. From these papers we identified subjects who were diagnosed with CTX, whose *CYP27A1* genotype and clinical case description was published in an individually identifying manner. For any subject that had been published in multiple papers, we reviewed and extracted clinical information about the subject from those papers as well, even if their date of publication fell outside of the range for the initial search, in order to compile the fullest clinical case description available in the literature. In addition, we identified functional data associated with pathogenic *CYP27A1* variants including at the level of mRNA, protein and enzyme acitivity.

### Data extraction

We extracted data on the age at onset, age at diagnosis, age at death, age of last report, age treatment started, treatment drug, cataract, optic disk pallor, tendon xanthomas, osteoporosis, osteopenia, pes cavus, intellectual disability, learning difficulty, developmental delay, cognitive decline, dementia, psychiatric disturbances, seizures, cerebellar signs, dysmetria, nystagmus, dysdiadochokinesia, dysarthria, speech disturbances, ataxia, gait abnormality, sensory ataxia, cerebellar atrophy, dentate nucleus lesions, cerebellar white matter lesions, intention tremor, pyramidal signs, weakness, hypotonia, spasticity, hyperreflexia, Babinski sign, spastic paraparesis, spastic tetraparesis, parkinsonism, resting tremor, postural tremor, bradykinesia, rigidity, hypomimia, dystonia, peripheral neuropathy, axonal neuropathy, demyelinating neuropathy, polyneuropathy, diarrhea, neonatal jaundice, and cholestasis. We also made a note of cholestanol levels, and any other biochemical testing results.

Cognition was a composite of intellectual disability, learning difficulty, dementia or cognitive decline; if an individual had any one of these features they were noted as having “cognition” to signify that CTX had affected their cognition.

Every paper was read at least twice, once each by two separate individuals.

Nomenclature for all variants follow HGVS standards and is reported in reference to CYP27A1 NM_000784.3 and NP_000775.1. Mutalyzer was used to generate HGVS nomenclature for all variants (89, 90).

### Genotype phenotype association

Individuals with CTX were divided into groups according to functional genotype. Those who had two LOF variants, defined as nonsense or frameshift variants, were grouped together and termed LOF. Those individuals who had two missense variants that were functionally demonstrated to have partial enzyme function were grouped together and referred to as hypomorphs. Individuals who had missense variants that were demonstrated to result in LOF were excluded from this analysis. Additionally, individuals who had splice variants were excluded from this analysis.

### Statistical analyses

The positive predictive value for each of the 12 most common clinical features of CTX was calculated as the number of individuals with the clinical feature divided by the number of individuals with the genotype of interest. The margin of error was calculated using t statistics.

## Author Contributions

PEB conceived and designed the study as well as analyzed and interpreted data. PEB wrote the article. JH extracted all of the data from the literature and helped draft the article.

## Acknowledgements

PEB is supported by NIH NINDS RO1 NS08372. The authors thank Dr. David Ledbetter for insightful conversation on IQ in Fragile X that directly influenced the thinking about IQ and cognition in CTX, discussed in this study.

## Data Availability

All data used in this study is publicly available in articles indexed in PubMed and in gnomAD https://gnomad.broadinstitute.org/.

## Human and Animal Subjects

This article does not contain any studies with human or animal subjects performed by any of the authors.

## References

1. Stelten BML, Huidekoper HH, van de Warrenburg BPC, Brilstra EH, Hollak CEM, Haak HR, et al. Long-term treatment effect in cerebrotendinous xanthomatosis depends on age at treatment start. Neurology. 2019;92(2):e83–e95.

2. Mignarri A, Gallus GN, Dotti MT, Federico A. A suspicion index for early diagnosis and treatment of cerebrotendinous xanthomatosis. J Inherit Metab Dis. 2014;37(3):421–9.

3. Amador MDM, Masingue M, Debs R, Lamari F, Perlbarg V, Roze E, et al. Treatment with chenodeoxycholic acid in cerebrotendinous xanthomatosis: clinical, neurophysiological, and quantitative brain structural outcomes. J Inherit Metab Dis. 2018;41(5):799–807.

4. Verrips A, Hoefsloot LH, Steenbergen GC, Theelen JP, Wevers RA, Gabreels FJ, et al. Clinical and molecular genetic characteristics of patients with cerebrotendinous xanthomatosis. Brain. 2000;123 (Pt 5):908–19.

5. Zádori D, Szpisjak L, Madar L, Varga VE, Csányi B, Bencsik K, et al. Different phenotypes in identical twins with cerebrotendinous xanthomatosis: case series. Neurol Sci. 2017;38(3):481–3.

6. Federico A, Dotti MT. Cerebrotendinous xanthomatosis: clinical manifestations, diagnostic criteria, pathogenesis, and therapy. J Child Neurol. 2003;18(9):633–8.

7. Moghadasian MH. Cerebrotendinous xanthomatosis: clinical course, genotypes and metabolic backgrounds. Clin Invest Med. 2004;27(1):42–50.

8. Leitersdorf E, Reshef A, Meiner V, Levitzki R, Schwartz SP, Dann EJ, et al. Frameshift and splice-junction mutations in the sterol 27-hydroxylase gene cause cerebrotendinous xanthomatosis in Jews or Moroccan origin. J Clin Invest. 1993;91(6):2488–96.

9. Reshef A, Meiner V, Berginer VM, Leitersdorf E. Molecular genetics of cerebrotendinous xanthomatosis in Jews of north African origin. J Lipid Res. 1994;35(3):478–83.

10. Leitersdorf E, Safadi R, Meiner V, Reshef A, Bjorkhem I, Friedlander Y, et al. Cerebrotendinous xanthomatosis in the Israeli Druze: molecular genetics and phenotypic characteristics. Am J Hum Genet. 1994;55(5):907–15.

11. Garuti R, Lelli N, Barozzini M, Dotti MT, Federico A, Bertolini S, et al. Partial deletion of the gene encoding sterol 27-hydroxylase in a subject with cerebrotendinous xanthomatosis. Journal of Lipid Research. 1996;37(3):662–72.

12. Garuti R, Lelli N, Barozzini M, Tiozzo R, Dotti MT, Federico A, et al. Cerebrotendinous xanthomatosis caused by two new mutations of the sterol-27-hydroxylase gene that disrupt mRNA splicing. Journal of Lipid Research. 1996;37(7):1459–67.

13. Garuti R, Croce MA, Tiozzo R, Dotti MT, Federico A, Bertolini S, et al. Four novel mutations of sterol 27-hydroxylase gene in Italian patients with cerebrotendinous xanthomatosis. Journal of Lipid Research. 1997;38(11):2322–34.

14. Brown AJ, Watts GF, Burnett JR, Dean RT, Jessup W. Sterol 27-hydroxylase acts on 7-ketocholesterol in human atherosclerotic lesions and macrophages in culture. J Biol Chem. 2000;275(36):27627–33.

15. Smalley SV, Preiss Y, Suazo J, Vega JA, Angellotti I, Lagos CF, et al. Novel splice-affecting variants in CYP27A1 gene in two Chilean patients with Cerebrotendinous Xanthomatosis. Genet Mol Biol. 2015;38(1):30–6.

16. Verrips A, Steenbergen-Spanjers GC, Luyten JA, Wevers RA, Wokke JH, Gabreels FJ, et al. Exon skipping in the sterol 27-hydroxylase gene leads to cerebrotendinous xanthomatosis. Hum Genet. 1997;100(2):284–6.

17. Chen W, Kubota S, Seyama Y. Alternative pre-mRNA splicing of the sterol 27-hydroxylase gene (CYP 27) caused by a G to A mutation at the last nucleotide of exon 6 in a patient with cerebrotendinous xanthomatosis (CTX). Journal of Lipid Research. 1998;39(3):509–17.

18. Chen W, Kubota S, Teramoto T, Nishimura Y, Yonemoto K, Seyama Y. Silent nucleotide substitution in the sterol 27-hydroxylase gene (CYP 27) leads to alternative pre-mRNA splicing by activating a cryptic 5’ splice site at the mutant codon in cerebrotendinous xanthomatosis patients. Biochemistry. 1998;37(13):4420–8.

19. Chen W, Kubota S, Ujike H, Ishihara T, Seyama Y. A novel Arg362Ser mutation in the sterol 27-hydroxylase gene (CYP27): its effects on pre-mRNA splicing and enzyme activity. Biochemistry. 1998;37(43):15050–6.

20. Chen W, Kubota S, Kim KS, Cheng J, Kuriyama M, Eggertsen G, et al. Novel homozygous and compound heterozygous mutations of sterol 27-hydroxylase gene (CYP27) cause cerebrotendinous xanthomatosis in three Japanese patients from two unrelated families. Journal of Lipid Research. 1997;38(5):870–9.

21. Chen W, Kubota S, Nishimura Y, Nozaki S, Yamashita S, Nakagawa T, et al. Genetic analysis of a Japanese cerebrotendinous xanthomatosis family: identification of a novel mutation in the adrenodoxin binding region of the CYP 27 gene. Biochim Biophys Acta. 1996;1317(2):119–26.

22. Kim KS, Kubota S, Kuriyama M, Fujiyama J, Björkhem I, Eggertsen G, et al. Identification of new mutations in sterol 27-hydroxylase gene in Japanese patients with cerebrotendinous xanthomatosis (CTX). Journal of Lipid Research. 1994;35(6):1031–9.

23. Zhang P, Zhao J, Peng XM, Qian YY, Zhao XM, Zhou WH, et al. Cholestasis as a dominating symptom of patients with CYP27A1 mutations: An analysis of 17 Chinese infants. J Clin Lipidol. 2021;15(1):116–23.

24. Sawada N, Sakaki T, Kitanaka S, Kato S, Inouye K. Structure-function analysis of CYP27B1 and CYP27A1. Studies on mutants from patients with vitamin D-dependent rickets type I (VDDR-I) and cerebrotendinous xanthomatosis (CTX). Eur J Biochem. 2001;268(24):6607–15.

25. Gupta RP, Patrick K, Bell NH. Mutational analysis of CYP27A1: assessment of 27-hydroxylation of cholesterol and 25-hydroxylation of vitamin D. Metabolism. 2007;56(9):1248–55.

26. Cali JJ, Hsieh CL, Francke U, Russell DW. Mutations in the bile acid biosynthetic enzyme sterol 27-hydroxylase underlie cerebrotendinous xanthomatosis. J Biol Chem. 1991;266(12):7779–83.

27. Kuwabara K, Hitoshi S, Nukina N, Ishii K, Momose T, Kubota S, et al. PET analysis of a case of cerebrotendinous xanthomatosis presenting hemiparkinsonism. J Neurol Sci. 1996;138(1-2):145–9.

28. Honda A, Salen G, Matsuzaki Y, Batta AK, Xu G, Leitersdorf E, et al. Side chain hydroxylations in bile acid biosynthesis catalyzed by CYP3A are markedly up-regulated in Cyp27-/-mice but not in cerebrotendinous xanthomatosis. J Biol Chem. 2001;276(37):34579–85.

29. Rystedt E, Olin M, Seyama Y, Buchmann M, Berstad A, Eggertsen G, et al. Cerebrotendinous xanthomatosis: molecular characterization of two Scandinavian sisters. J Intern Med. 2002;252(3):259–64.

30. Hoflinger P, Hauser S, Yutuc E, Hengel H, Griffiths L, Radelfahr F, et al. Metabolic profiling in serum, cerebrospinal fluid, and brain of patients with cerebrotendinous xanthomatosis. J Lipid Res. 2021;62:100078.

31. Chang WN, Kuriyama M, Lui CC, Jeng SF, Chen WJ, Chee EC. Cerebrotendinous xanthomatosis in three siblings from a Taiwanese family. J Formos Med Assoc. 1992;91(12):1190–4.

32. Lee MJ, Huang YC, Sweeney MG, Wood NW, Reilly MM, Yip PK. Mutation of the sterol 27-hydroxylase gene (CYP27A1) in a Taiwanese family with cerebrotendinous xanthomatosis. J Neurol. 2002;249(9):1311–2.

33. Wang PW, Chang WN, Lu CH, Chao D, Schrag C, Pan TL. New insights into the pathological mechanisms of cerebrotendinous xanthomatosis in the Taiwanese using genomic and proteomic tools. Proteomics. 2006;6(3):1029–37.

34. Su CS, Chang WN, Huang SH, Lui CC, Pan TL, Lu CH, et al. Cerebrotendinous xanthomatosis patients with and without parkinsonism: clinical characteristics and neuroimaging findings. Mov Disord. 2010;25(4):452–8.

35. Chang CC, Lui CC, Wang JJ, Huang SH, Lu CH, Chen C, et al. Multi-parametric neuroimaging evaluation of cerebrotendinous xanthomatosis and its correlation with neuropsychological presentations. BMC Neurol. 2010;10:59.

36. Chen SF, Tsai NW, Chang CC, Lu CH, Huang CR, Chuang YC, et al. Neuromuscular abnormality and autonomic dysfunction in patients with cerebrotendinous xanthomatosis. BMC Neurol. 2011;11:63.

37. Chen SF, Chang CC, Huang SH, Lu CH, Chuang YC, Pan TL, et al. 99mTc-sestamibi thigh SPECT/CT imaging for assessment of myopathy in cerebrotendinous xanthomatosis with histopathological and immunohistochemical correlation. Clin Nucl Med. 2014;39(3):e202–7.

38. Zhang S, Li W, Zheng R, Zhao B, Zhang Y, Zhao D, et al. Cerebrotendinous xanthomatosis with peripheral neuropathy: a clinical and neurophysiological study in Chinese population. Ann Transl Med. 2020;8(21):1372.

39. Zadori D, Szpisjak L, Madar L, Varga VE, Csanyi B, Bencsik K, et al. Different phenotypes in identical twins with cerebrotendinous xanthomatosis: case series. Neurol Sci. 2017;38(3):481–3.

40. Giraldo-Chica M, Acosta-Baena N, Urbano L, Velilla L, Lopera F, Pineda N. Novel cerebrotendinous xanthomatosis mutation causes familial early dementia in Colombia. Biomedica. 2015;35(4):563–71.

41. Yunisova G, Tufekcioglu Z, Dogu O, Bilgic B, Kaleagasi H, Akca Kalem S, et al. Patients with Lately Diagnosed Cerebrotendinous Xanthomatosis. Neurodegener Dis. 2019;19(5-6):218–24.

42. Wakamatsu N, Hayashi M, Kawai H, Kondo H, Gotoda Y, Nishida Y, et al. Mutations producing premature termination of translation and an amino acid substitution in the sterol 27-hydroxylase gene cause cerebrotendinous xanthomatosis associated with parkinsonism. J Neurol Neurosurg Psychiatry. 1999;67(2):195–8.

43. Kisa PT, Yildirim GK, Hismi BO, Dorum S, Kusbeci OY, Topak A, et al. Patients with cerebrotendinous xanthomatosis diagnosed with diverse multisystem involvement. Metab Brain Dis. 2021;36(6):1201–11.

44. Posada IJ, Ramos A. Botulinum toxin-responsive oromandibular dystonia in cerebrotendinous xanthomatosis. Parkinsonism Relat Disord. 2011;17(7):570–2.

45. Schotsmans K, De Cauwer H, Baets J, Ceyssens S, van den Hauwe L, Deconinck T, et al. Cerebrotendinous xanthomatosis presenting with asymmetric parkinsonism: a case with I-123-FP-CIT SPECT imaging. Acta Neurol Belg. 2012;112(3):287–9.

46. Pilo-de-la-Fuente B, Jimenez-Escrig A, Lorenzo JR, Pardo J, Arias M, Ares-Luque A, et al. Cerebrotendinous xanthomatosis in Spain: clinical, prognostic, and genetic survey. Eur J Neurol. 2011;18(10):1203–11.

47. Pilo de la Fuente B, Sobrido MJ, Girós M, Pozo L, Lustres M, Barrero F, et al. Usefulness of cholestanol levels in the diagnosis and follow-up of patients with cerebrotendinous xanthomatosis. Neurología (English Edition). 2011;26(7):397–404.

48. Pilo de la Fuente B, Ruiz I, Lopez de Munain A, Jimenez-Escrig A. Cerebrotendinous xanthomatosis: neuropathological findings. J Neurol. 2008;255(6):839–42.

49. Pilo B, de Blas G, Sobrido MJ, Navarro C, Grandas F, Barrero FJ, et al. Neurophysiological study in cerebrotendinous xanthomatosis. Muscle Nerve. 2011;43(4):531–6.

50. Mignarri A, Dotti MT, Federico A, De Stefano N, Battaglini M, Grazzini I, et al. The spectrum of magnetic resonance findings in cerebrotendinous xanthomatosis: redefinition and evidence of new markers of disease progression. J Neurol. 2017;264(5):862–74.

51. Dotti MT, Salen G, Federico A. Cerebrotendinous xanthomatosis as a multisystem disease mimicking premature ageing. Dev Neurosci. 1991;13(4-5):371–6.

52. Mondelli M, Rossi A, Scarpini C, Dotti MT, Federico A. Evoked potentials in cerebrotendinous xanthomatosis and effect induced by chenodeoxycholic acid. Arch Neurol. 1992;49(5):469–75.

53. Dotti MT, Rufa A, Federico A. Cerebrotendinous xanthomatosis: heterogeneity of clinical phenotype with evidence of previously undescribed ophthalmological findings. J Inherit Metab Dis. 2001;24(7):696–706.

54. Mignarri A, Rossi S, Ballerini M, Gallus GN, Del Puppo M, Galluzzi P, et al. Clinical relevance and neurophysiological correlates of spasticity in cerebrotendinous xanthomatosis. J Neurol. 2011;258(5):783–90.

55. Ginanneschi F, Mignarri A, Mondelli M, Gallus GN, Del Puppo M, Giorgi S, et al. Polyneuropathy in cerebrotendinous xanthomatosis and response to treatment with chenodeoxycholic acid. J Neurol. 2013;260(1):268–74.

56. Roeben B, Just J, Hengel H, Bender F, Poschl P, Synofzik M, et al. Multifocal, hypoechogenic nerve thickening in Cerebrotendinous Xanthomatosis. Clin Neurophysiol. 2020;131(8):1798–803.

57. Stelten BML, Lycklama ANGJ, Hendriks E, Kluijtmans LAJ, Wevers RA, Verrips A. Long-term MRI Findings in Patients With Cerebrotendinous Xanthomatosis Treated With Chenodeoxycholic Acid. Neurology. 2022;99(13):559–66.

58. Kim S, Park JS, Lee JH, Shin HY, Yang HJ, Shin JH. Clinical, electrophysiological, and genetic characteristics of cerebrotendinous xanthomatosis in South Korea. Neurocase. 2022;28(6):477–82.

59. Li J, Xu EH, Mao W, Qiao HW, Zhou YT, Yang Q, et al. Parkinsonism with Normal Dopaminergic Presynaptic Terminals in Cerebrotendinous Xanthomatosis. Mov Disord Clin Pract. 2020;7(1):115–6.

60. Stelten BML, Bonnot O, Huidekoper HH, van Spronsen FJ, van Hasselt PM, Kluijtmans LAJ, et al. Autism spectrum disorder: an early and frequent feature in cerebrotendinous xanthomatosis. J Inherit Metab Dis. 2018;41(4):641–6.

61. Mignarri A, Falcini M, Vella A, Giorgio A, Gallus GN, Del Puppo M, et al. Parkinsonism as neurological presentation of late-onset cerebrotendinous xanthomatosis. Parkinsonism Relat Disord. 2012;18(1):99–101.

62. Dotti MT, Federico A, Garuti R, Calandra S. Cerebrotendinous xanthomatosis with predominant parkinsonian syndrome: further confirmation of the clinical heterogeneity. Mov Disord. 2000;15(5):1017–9.

63. Nakamura S, Tamura T, Takahashi H, Ishida-Yamamoto A, Hashimoto Y, Kuroda K, et al. Cerebrotendinous xanthomatosis: report of a case. Br J Dermatol. 2000;142(2):378–80.

64. Lee CW, Lee JJ, Lee YF, Wang PW, Pan TL, Chang WN, et al. Clinical and molecular genetic features of cerebrotendinous xanthomatosis in Taiwan: Report of a novel CYP27A1 mutation and literature review. J Clin Lipidol. 2019;13(6):954–9 e1.

65. Yu T, Nie S, Bu L, Liu M, He J, Niu X, et al. Cholestanol accelerates alpha-synuclein aggregation and spreading by activating asparagine endopeptidase. JCI Insight. 2023;8(21).

66. DeBarber AE, Luo J, Giugliani R, Souza CF, Chiang JP, Merkens LS, et al. A useful multi-analyte blood test for cerebrotendinous xanthomatosis. Clin Biochem. 2014;47(9):860–3.

67. DeBarber AE, Connor WE, Pappu AS, Merkens LS, Steiner RD. ESI-MS/MS quantification of 7alpha-hydroxy-4-cholesten-3-one facilitates rapid, convenient diagnostic testing for cerebrotendinous xanthomatosis. Clin Chim Acta. 2010;411(1-2):43–8.

68. DeBarber AE, Kalfon L, Fedida A, Fleisher Sheffer V, Ben Haroush S, Chasnyk N, et al. Newborn screening for cerebrotendinous xanthomatosis is the solution for early identification and treatment. J Lipid Res. 2018;59(11):2214–22.

69. DeBarber AE, Schaefer EJ, Do J, Ray JW, Larson A, Redder S, et al. Genetically and clinically confirmed atypical cerebrotendinous xanthomatosis with normal cholestanol and marked elevations of bile acid precursors and bile alcohols. Journal of Clinical Lipidology. 2024.

70. Vega GL, Illingworth DR, Grundy SM, Lindgren FT, Connor WE. Normocholesterolemic tendon xanthomatosis with overproduction of apolipoprotein B. Metabolism. 1983;32(2):118–25.

71. Guenzel AJ, DeBarber A, Raymond K, Dhamija R. Familial variability of cerebrotendinous xanthomatosis lacking typical biochemical findings. JIMD Reports. 2021;59(1):3–9.

72. Valencia-Sanchez C, Wingerchuk DM, Dhamija R. Teaching NeuroImages: Spinal xanthomatosis: A misdiagnosed, treatable cause of progressive myelopathy. Neurology. 2020;95(11):e1615–e6.

73. Wallon D, Guyant-Marechal L, Laquerriere A, Wevers RA, Martinaud O, Kluijtmans LA, et al. Clinical imaging and neuropathological correlations in an unusual case of cerebrotendinous xanthomatosis. Clin Neuropathol. 2010;29(6):361–4.

74. Bonnet JB, Couvert P, Di-Filippo M, Boissiere F, Cristol JP, Sutra T, et al. Tuberous xanthomatosis is not necessarily associated with increased plasma concentrations of cholestanol in cerebrotendinous xanthomatosis. J Intern Med. 2023;293(1):121–3.

75. Stelten BML, Raal FJ, Marais AD, Riksen NP, Roeters van Lennep JE, Duell PB, et al. Cerebrotendinous xanthomatosis without neurological involvement. J Intern Med. 2021;290(5):1039–47.

76. Takahashi M, Okazaki H, Tada H, Ishibashi S. A case of cerebrotendinous xanthomatosis with massive xanthomas but without a considerable increase in serum cholestanol levels. J Clin Lipidol. 2023;17(6):834–8.

77. Kapas I, Katko M, Harangi M, Paragh G, Balogh I, Koczi Z, et al. Cerebrotendinous xanthomatosis with the c.379C>T (p.R127W) mutation in the CYP27A1 gene associated with premature age-associated limbic tauopathy. Neuropathol Appl Neurobiol. 2014;40(3):345–50.

78. Chen C, Zhang Y, Wu H, Sun YM, Cai YH, Wu JJ, et al. Clinical and molecular genetic features of cerebrotendinous xanthomatosis patients in Chinese families. Metab Brain Dis. 2017;32(5):1609–18.

79. Tao QQ, Zhang Y, Lin HX, Dong HL, Ni W, Wu ZY. Clinical and genetic characteristics of Chinese patients with cerebrotendinous xanthomatosis. Orphanet J Rare Dis. 2019;14(1):282.

80. Wang Z, Yuan Y, Zhang W, Zhang Y, Feng L. Cerebrotendinous xanthomatosis with a compound heterozygote mutation and severe polyneuropathy. Neuropathology. 2007;27(1):62–6.

81. Tibrewal S, Duell PB, DeBarber AE, Loh AR. Cerebrotendinous xanthomatosis: early diagnosis on the basis of juvenile cataracts. J AAPOS. 2017;21(6):505–7.

82. Duell PB, Salen G, Eichler FS, DeBarber AE, Connor SL, Casaday L, et al. Diagnosis, treatment, and clinical outcomes in 43 cases with cerebrotendinous xanthomatosis. J Clin Lipidol. 2018;12(5):1169–78.

83. Verrips A, Nijeholt GJ, Barkhof F, Van Engelen BG, Wesseling P, Luyten JA, et al. Spinal xanthomatosis: a variant of cerebrotendinous xanthomatosis. Brain. 1999;122 (Pt 8):1589–95.

84. Takasone K, Morizumi T, Nakamura K, Mochizuki Y, Yoshinaga T, Koyama S, et al. A Late-onset and Relatively Rapidly Progressive Case of Pure Spinal Form Cerebrotendinous Xanthomatosis with a Novel Mutation in the CYP27A1 Gene. Intern Med. 2020;59(20):2587–91.

85. Cali JJ, Russell DW. Characterization of human sterol 27-hydroxylase. A mitochondrial cytochrome P-450 that catalyzes multiple oxidation reaction in bile acid biosynthesis. J Biol Chem. 1991;266(12):7774–8.

86. Prosser DE, Guo Y, Jia Z, Jones G. Structural motif-based homology modeling of CYP27A1 and site-directed mutational analyses affecting vitamin D hydroxylation. Biophys J. 2006;90(10):3389–409.

87. Bhattacharyya AK, Lin DS, Connor WE. Cholestanol metabolism in patients with cerebrotendinous xanthomatosis: absorption, turnover, and tissue deposition. J Lipid Res. 2007;48(1):185–92.

88. Lutjohann D, Stellaard F, Bjorkhem I. Levels of 7alpha-hydroxycholesterol and/or 7alpha-hydroxy-4-cholest-3-one are the optimal biochemical markers for the evaluation of treatment of cerebrotendinous xanthomatosis. J Neurol. 2020;267(2):572–3.

89. Lefter M, Vis JK, Vermaat M, den Dunnen JT, Taschner PEM, Laros JFJ. Mutalyzer 2: next generation HGVS nomenclature checker. Bioinformatics. 2021;37(18):2811–7.

90. Vis JK, Vermaat M, Taschner PE, Kok JN, Laros JF. An efficient algorithm for the extraction of HGVS variant descriptions from sequences. Bioinformatics. 2015;31(23):3751–7.

